# Determinants of virological non-suppression among children receiving antiretroviral therapy in Chókwè, Mozambique: a retrospective cohort study

**DOI:** 10.64898/2026.07.21.26358610

**Authors:** Félix Fane Cubai, Raquel de Vasconcellos Carvalhaes de Oliveira, Igor Paulo Ubisse Capitine, Andrey Moreira Cardoso

## Abstract

**Introduction:** Despite major progress in antiretroviral therapy (ART) scale-up, viral suppression (VS) among children living with Human Immunodeficiency Virus (HIV) remains below global targets in Mozambique. Evidence on determinants of virological non-suppression (VNS) in decentralized rural settings is still limited

**Objective:** This study aimed to analyse factors associated with VNS among children receiving ART in Chókwè District, Gaza Province, Mozambique.

**Methods:** We conducted a retrospective cohort study including children younger than 15 years who initiated ART between 2022 and 2024 in 27 health facilities in Chókwè District. Sociodemographic, clinical, laboratory, and health care data were extracted from paper medical records and electronic databases. VNS was defined as viral load (VL) ≥1,000 copies/mL of blood after at least 6 months of ART initiation. Kaplan–Meier methods and Cox proportional hazards regression models, adjusted for key sociodemographic, clinical, and health care characteristics, were used to identify factors associated with time to VNS. Adjusted hazard ratios (aHR) and 95% confidence intervals (95% CI) were estimated.We conducted statistical analyses using the STATA 15 and R 4.5.1 software packages.

**Results:** A total of 285 children were included. At 6 months after ART initiation, 28.2% of children presented VNS, while 23.3% presented VNS at 18 months. In the adjusted analysis for the first 6 months of follow-up each additional year of age at ART initiation was associated with lower risk of VNS, whereas not initiating treatment on the same day of diagnosis increased the risk of VNS. At 18 months of follow-up, age at ART initiation showed evidence of a non-linear association with VNS. Tuberculosis co-infection and undernutrition at 6 months were also associated with a higher risk of VNS.

**Conclusion:** VNS remains frequent among children receiving ART in Chókwè District. Younger age during early follow-up, delayed ART initiation, tuberculosis co-infection, and undernutrition were important determinants of poor virological outcomes. Strengthening early HIV diagnosis, timely ART initiation, integrated management of TB and undernutrition, and age-responsive follow-up strategies may improve long-term virological outcomes among children living with HIV in resource-limited settings.

## Introduction

Human Immunodeficiency Virus (HIV) infection remains a major global public health challenge, particularly in sub-Saharan Africa, where children continue to experience lower rates of viral suppression (VS) compared to adults. Despite substantial progress in antiretroviral therapy (ART) scale-up, achieving optimal virological outcomes among children living with HIV remains difficult in many resource-limited settings (1). In 2024, an estimated 1.4 million children younger than 15 years were living with HIV globally, with the majority residing in Eastern and Southern Africa (1). In Mozambique, according to the latest National Survey on the Impact of HIV and AIDS (INSIDA 2021), approximately 2.4 million people were living with HIV, with a prevalence of 12.5% among individuals aged 15 years and older. No prevalence were estimated for individuals younger than 15 years (2).

The UNAIDS 95-95-95 strategy emphasizes the importance of universal HIV diagnosis, ART coverage, and VS to end the HIV epidemic as a public health threat by 2030 (3). Although major progress has been achieved globally, children remain disproportionately affected by poorer treatment outcomes and lower rates of VS compared to adults (1). Viral load (VL) monitoring is currently the recommended standard for assessing treatment response and identifying treatment failure in people receiving ART (4,5).

In Mozambique, routine VL monitoring has been progressively expanded since 2015 and currently represents the main strategy for evaluating ART response and identifying treatment failure (6).

According to national guidelines, VL testing is recommended six months after ART initiation and annually thereafter in patients with VS (6). Children with VL ≥1,000 copies/mL are referred for enhanced adherence counseling (EAC), consisting of structured adherence support sessions followed by repeat VL testing (6).

Despite improvements in ART access and VL monitoring, important inequalities persist across age groups in Mozambique. Children younger than 15 years remain among the populations with the lowest VS rates nationally (7). In Gaza Province and particularly in Chókwè District, pediatric HIV care continues to face important challenges related to treatment adherence, long-term follow-up, retention in care, and virological monitoring.

Key challenges to achieve treatment success include difficulties with treatment adherence, late diagnosis, prior exposure to antiretrovirals (ARVs), limited diagnostic capacity, shortages of human resources, challenges in longitudinal follow-up, and socio-familial factors (8,9). Previous studies have identified several factors associated with VNS among children receiving ART, including poor adherence, delayed diagnosis, opportunistic infections, prior exposure to ARVs, advanced HIV disease, treatment regimen changes, and psychosocial barriers affecting caregivers and children (8–11). Structural barriers such as stigma, food insecurity, transportation difficulties, and weak family support may further compromise treatment continuity and adherence. In addition, tuberculosis (TB) co-infection remains an important challenge in pediatric HIV care, particularly in high-burden settings in sub-Saharan Africa.

Evidence on determinants of VNS among children receiving ART in decentralized and rural settings in Mozambique remains limited, particularly using longitudinal analyses integrating sociodemographic, clinical, and health care factors. This lack of context-specific evidence constrains the development of targeted interventions to improve pediatric HIV outcomes in resource-limited settings.

This study aimed to identify determinants of VNS among children younger than 15 years receiving ART in Chókwè District, Mozambique, using longitudinal programmatic data from decentralized health facilities. We hypothesized that younger age, delayed ART initiation and TB co-infection would be associated with increased risk of VNS.

## Materials and methods

### Design

This retrospective cohort study was conducted to identify determinants of VNS in children under 15 years who initiated ART for HIV between January 2022 and December 2024 in the Chókwè District, Gaza Province – Mozambique – The Chókwè Cohort Study (CCS). The study period was defined to start in the post–COVID-19 moment in which health services and pediatric VL monitoring progressively resumed. Data collection ended in November 2025, allowing a minimum potential follow-up period of 18 months.

### Study setting

Mozambique introduced antiretroviral therapy (ART) gradually in 2003 and has made significant progress in its response over the years, with a greater focus on improving access to ART and prevention services. Chókwè District is in Gaza Province, southern Mozambique (Fig 1). The district covers approximately 2,600 km² and has an estimated population of 246.617 inhabitants (2026 projection), accounting for approximately 18% of the province’s total population, with a population density of 92 inhabitants per km² (12). HIV seroprevalence in the district is estimated at 24.1% (2).

**Fig 1.**
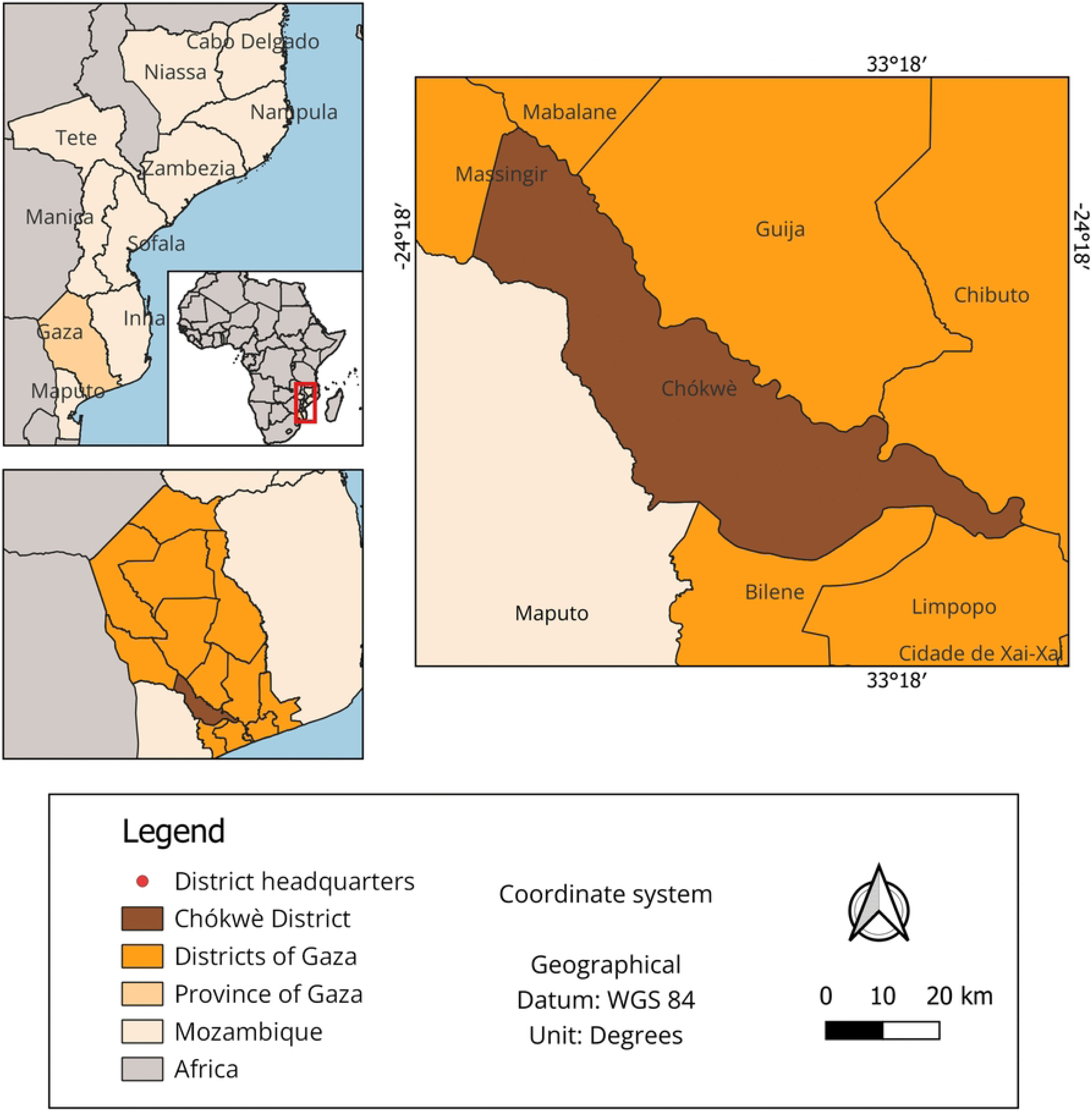
Geographical location of the Chókwè district, Gaza Province, Mozambique. Chókwè Cohort Study, 2022-2024. Map illustrating the location of Chókwè District within Gaza Province and Mozambique. The figure provides geographical context for the study area, indicating district boundaries and its position in southern Mozambique. Map elements include orientation and scale.

Between 2022 and 2024, Chókwè District had 27 health facilities providing ART services, all included in this study. According to the National Health Information System for Monitoring and Evaluation (SIS-MA) data, by December 2024 the district had 3,028 active patients on ART, including 359 (11.9%) children aged <15 years, highlighting the relevance of pediatric HIV care in the district. SIS-MA, built on the District Health Information Software 2 (DHIS2), is the national platform used in Mozambique for routine collection, management, and monitoring of health data to support health program evaluation and decision-making (13).

### Population

A total of 285 eligible children were enrolled in the cohort. Children without viral load measurements were not considered to have experienced the study outcome; instead, they contributed follow-up time until their last documented clinical contact and were subsequently treated as censored in the survival analysesThe study included all children who met both the inclusion and exclusion criteria. No sampling procedures were performed. Instead, a census approach was used. The inclusion criteria was defined as children aged <15 years who initiated ART between January 2022 and December 2024 in the 27 health facilities providing ART services in Chókwè District, with available records in patients’ paper charts or in the Determined, Resilient, Empowered, AIDS-free, Mentored and Safe (DREAMS) Layering Tool electronic system. The exclusion criteria included children who initiated ART outside Chókwè District. Additionally, for the analysis of VNS at 6 and 18 months, children who were transferred out of the district before completing follow-up and those without VL results during the follow-up periods were excluded, totaling 285 patients.

### HIV diagnosis and clinical management

In Chókwè district, children are managed according to the Mozambican national HIV care guidelines, which are aligned with World Health Organization (WHO) recommendations and constitute the standard reference for HIV management in public health facilities (14).

HIV testing for children <15 years is initiated based on clinical suspicion, parental HIV status, or through provider-initiated testing and counselling (PITC) at outpatient, inpatient, or Prevention of Mother-To-Child Transmission (PMTCT) services. Laboratory diagnostic confirmation is performed by a rapid diagnostic test algorithm for children ≥18 months or by polymerase chain reaction (PCR) testing, for those <18 months. All confirmed HIV-positive children are enrolled in ART care and screened for opportunistic infections, including tuberculosis (TB) (14).

TB screening is performed at every visit using the national TB symptom checklist (cough, fever, weight loss, or night sweats). In children, particular attention is given to growth faltering and history of contact with TB cases. Those with presumptive TB were further investigated using GeneXpert MTB/RIF and chest X-ray, in accordance with national guidelines (14).

First-line ART for children consist of a Dolutegravir (DTG)**-**based regimen, using ABC/3TC **+** DTG or TDF/3TC **+** DTG, depending on the child’s weight and age. Clinicians prescribe second-line ART for patients experiencing treatment failure, depending on first line regimen used. Possible combinations included AZT/3TC **+** LPV/r or ATV/r, AZT/3TC + DTG, TDF/3TC **+** DTG, or ABC/3TC **+** DTG, following the national HIV infection management in children and adolescents’ treatment algorithms. ART initiation occurs on the same day as HIV diagnosis whenever possible. Routine clinical and laboratory monitoring includes CD4 cell count and VL testing (8).

### Viral load count

HIV-1 VL count is routinely performed in the reference laboratory using the Hologic Panther automated platform via the Aptima HIV-1 Quant Dx assay. The test is based on Transcription-Mediated Amplification (TMA) technology, which allows isothermal amplification of viral RNA after an automated nucleic acid capture and extraction step.

The assay is primarily validated for plasma samples, considered the gold standard for VL monitoring, using an input volume of 0.5 mL. However, it can also be applied to dried blood spots (DBS) samples, especially in programmatic or resource-limited settings where plasma processing and transport may be difficult. Evaluation studies demonstrate that the method has a limit of detection (LoD) of approximately 12 copies/mL and a lower limit of quantification (LLoQ) of 30 copies/mL, with a wide linear range of quantification (15).

HIV-1 viral load testing was performed using the Aptima HIV-1 Quant Dx assay (Hologic Panther platform), a validated automated molecular assay routinely used for HIV monitoring in Mozambique (15,16).

In the present study, we used only the VL results recorded in the laboratory system, with no intervention by the researchers in the sample collection or processing stages. The determinations followed the laboratory’s standard operating procedures and national guidelines for monitoring virological response in patients on ART.

### Psychological support

All children and caregivers receive psychosocial counseling focused on adherence, stigma reduction, and family engagement. We provide additional support to participants facing barriers such as food insecurity, transport difficulties, or family instability in accordance with the National Guideline for Implementation of Health Counseling and Testing in Mozambique (17).

### HIV diagnosis disclosure

Disclosing an HIV diagnosis is the process of informing a child of their serological status, comprising two phases: 1) Partial disclosure – when we discuss the disease without mentioning its name; 2) Full disclosure – when the name of the disease is mentioned (17).

### Conceptual framework and study variables

A conceptual framework outlining the determinants of VNS among children receiving ART guided the selection of explanatory variables (Fig 2). This framework incorporates three main dimensions (sociodemographic, clinical-laboratory, and health care) which, according to the literature, influence this outcome. We used standardized operational definitions to ensure comparability and consistency of the analysed data for this study.

**Fig 2.**
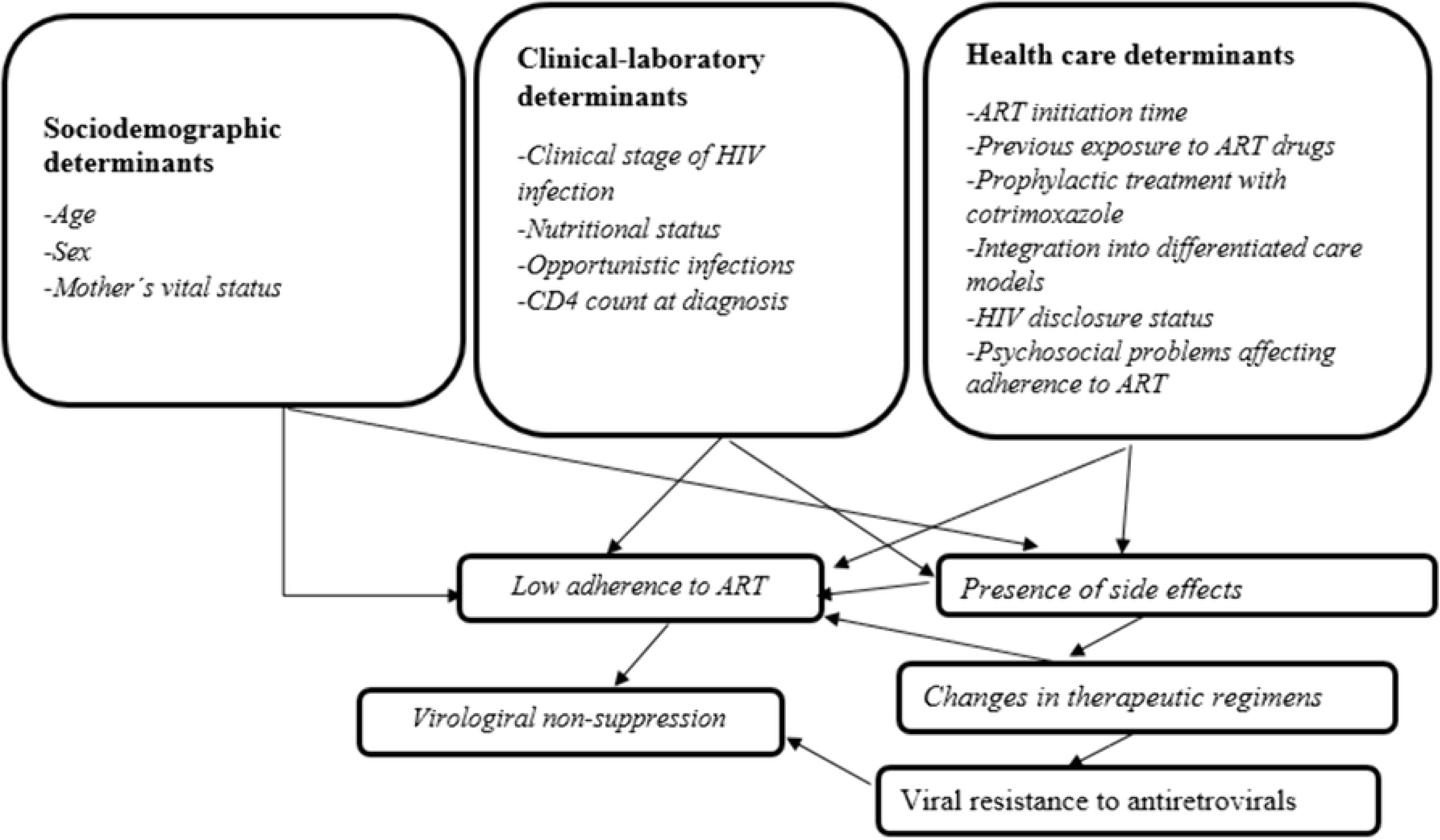
Theoretical model for determinants of virological non-suppression in children on ART. Chókwè Cohort Study, 2022-2024. Conceptual framework illustrating the relationships between sociodemographic, clinical–laboratory, and health care determinants associated with VNS among children receiving ART in Chókwè District, Mozambique, between 2022 and 2024. The model is structured to reflect proximal and distal factors influencing virological outcomes over time. ART: antiretroviral therapy; VNS: Virological non-suppression; WHO: World Health Organization; CD4: cluster of differentiation 4.

### Explanatory variables

Explanatory variables were grouped into three domains: sociodemographic, clinical–laboratory, and health care characteristics, based on the conceptual framework and previous literature on determinants of virological outcomes in pediatric HIV care. Variables included age, sex, maternal vital status, WHO clinical stage, nutritional status, TB co-infection, CD4 count, ART-related characteristics, adherence indicators, disclosure of HIV status, psychosocial barriers, and psychological support. Some variables were measured at baseline, whereas others were assessed during the 6-month follow-up period. Detailed operational definitions and categorizations of variables are presented in S1 Table.

### Follow-up, outcome, and censoring variables

#### Follow-up

Assessments were conducted at 6 months (defined as 180 days after ART initiation) and 18 months (defined as 540 days after ART initiation). Recognizing that VL measurements are not always performed exactly at the scheduled time points due to operational and logistical constraints, a tolerance window of up to three months was applied for each follow-up assessment. Accordingly, VL measurements were considered valid if performed between 180 and 270 days for the 6-month assessment, and between 540 and 630 days for the 18-month assessment.

#### Outcome

Virological non-suppression (VNS), defined as VL ≥1000 copies/ml of blood after at least six months on ART. The viral suppression (VS) was defined as VL <1000 copies/ml of blood, consistent with national and WHO guidelines (6).

#### Censoring

children who remained active on ART at the end of follow-up, transferred out, lost follow-up, or died before completing the expected follow-up period were considered censored in time-based analyses.

Loss of follow-up (LTFU) was defined as a child who missed scheduled ART refill or clinical appointments for more than 60 days after the last expected visit and was not confirmed as dead or transferred out. Even though their final outcome was unknown, these individuals still contributed “person-time” to the study up until the last point they were successfully followed; transfer out, as a child formally transferred to another ART facility with documented transfer in the patients’ paper charts or electronic medical records; and death as a documented death during the follow-up period.

### Data sources and management

Data were obtained from patients’ paper charts and from electronic medical records extracted via the DREAMS Layering Tool (DLT). Developed within the DREAMS initiative, the DLT is an electronic data management system designed to capture and track individual-level receipt of multiple interventions (“layering”) among adolescent girls and young women, supporting routine monitoring and evaluation of program implementation (18).

In the study setting, the system is implemented in two health facilities - Chalucuane Health Center and Carmelo Health Center - and captures the same variables routinely recorded in patients’ paper charts, including clinical, demographic, and laboratory information. We also reviewed clinical and laboratory registers to verify and validate the collected data. Where we identified discrepancies, we cross-checked against primary source documents.

Trained research assistants, under the supervision of the principal investigator, conducted the data collection. All data were anonymized and entered into Microsoft Excel to create the study database. Data confidentiality was ensured by storing the electronic database on password-protected computers and keeping physical documents in a secure location accessible only to the research team.

The research team accessed the medical records and electronic databases for data extraction between August 20, 2025 and November 30, 2025.

### Statistical analysis

Descriptive statistics were used to characterize the study population. We summarized categorical variables using absolute and relative frequencies, and continuous variables were described using measures of central tendency and dispersion.

We performed two time-to-event analyses using survival analysis, considering the time since ART initiation until the occurrence of VNS (at 6 months and 18 months) as the dependent variables (outcomes). The first event was a VL ≥ 1.000 copies/mL at 6 months of ART, and the second event was a VL ≥ 1.000 copies/mL at 18 months of ART. We censored patients who were followed up to the outcome assessment point with a VL < 1,000 copies/mL using the date of VL testing, or those who dropped out of treatment (including deaths, missing VL counts, or transfers outside the district), using the date of last contact or available information.

We used Kaplan–Meier method to estimate the cumulative probability of remaining free from VNS over time. Differences between groups were assessed using Kaplan–Meier curves and compared using the log-rank test (19). To identify factors associated with time to VNS, Cox proportional hazards regression models were fitted, with results expressed as crude and adjusted hazard ratios (HR) and corresponding 95% confidence intervals (95% CI) (20). We conducted analyses separately for two follow-up periods: up to 6 months after ART initiation and from 0 to 18 months of follow-up.

We initially fitted single-covariate Cox regression models for each covariate. Variables included in the multi-covariate Cox regression models were selected a priori based on epidemiological relevance, as informed by previous literature and the study conceptual framework, rather than solely on statistical significance in univariable analyses. For the 6-month VNS outcome, the variables considered were sex, age at ART initiation, maternal vital status, nutritional status at ART initiation, same-day as diagnosis ART initiation, cotrimoxazole preventive therapy at ART initiation, and TB co-infection at ART initiation. For the 18-month VNS outcomes, the variables considered were sex, age at ART initiation, maternal vital status, same-day as diagnosis ART initiation, cotrimoxazole preventive therapy at 6 months, nutritional status at 6 months, TB at 6 months, psychological support, differentiated care models, adherence to ART, and disclosure of HIV status. To control for baseline risk differences, multicovariate Cox models for the 6 and 18-month periods of ART were stratified by baseline clinical stage of HIV infection.

We initially assessed age at ART initiation for its functional form using penalized splines. In the model for the first 6 months of follow-up, age exhibited linear behavior, so we included it as a continuous variable in the final model. In contrast, the 18-month ART model shows a non-linear effect of age, so we modeled age using penalized splines in the final model.

Multicollinearity among covariates was assessed using the generalized variance inflation factor (GVIF) and found no evidence of either multicollinearity in the 6-month ART or the 18-month ART models (GVIF adjusted range: 1.00–1.55). Schoenfeld residuals confirmed the proportional hazards assumption in either the 6-month or 18-month models. We adopted a two-sided significance level of 5% (α = 0.05) for all analyses.

All analyses were conducted using STATA version 15 and R version 4.5.1 (21). The following packages were used: survival (22) and survminer (23).

### Ethical considerations

The National Bioethics Committee for Health of Mozambique (CNBS) approved the study protocol on August 19, 2025, under reference number 486/CNBS/2025. We obtained prior authorizations from the Research Ethics Committee of the Sergio Arouca National School of Public Health, Oswaldo Cruz Foundation, Brazil and the National Institute of Health (INS), Mozambique. The Provincial Health Directorate of Gaza and the District Health Services of Chókwè granted additional approval.

Because this study relied exclusively on secondary data from patients’ paper medical records and the DREAMS Layering Toll system, we could not obtain individual informed consent retrospectively. However, we obtained formal authorization from the institutions responsible for data custody and access.

To ensure participant privacy, all patients’ paper medical records were anonymized, with no collection of personal identifiers. Electronic databases were stored on password-protected computers with access restricted to the research team, and physical documents were kept in a secure location.

## Results

### Viral load cascade and proportion of outcomes

The participant selection process, the VL cascade, and the proportion of outcome in children enrolled for the study are presented in Fig 3. At 6 months of ART, 285 children were included, of whom 227 had VL results and 58 were censored for various reasons. Among those with VL results, 163 (71.8%) children achieved VS while 64 (28.2%) had VNS. Between 6 and 18 months of ART, 129 children had VL results, and 98 were censored. Among those with VL results, 99 (76.7%) achieved VS, whereas 30 (23.3%) had VNS.

**Fig 3.**
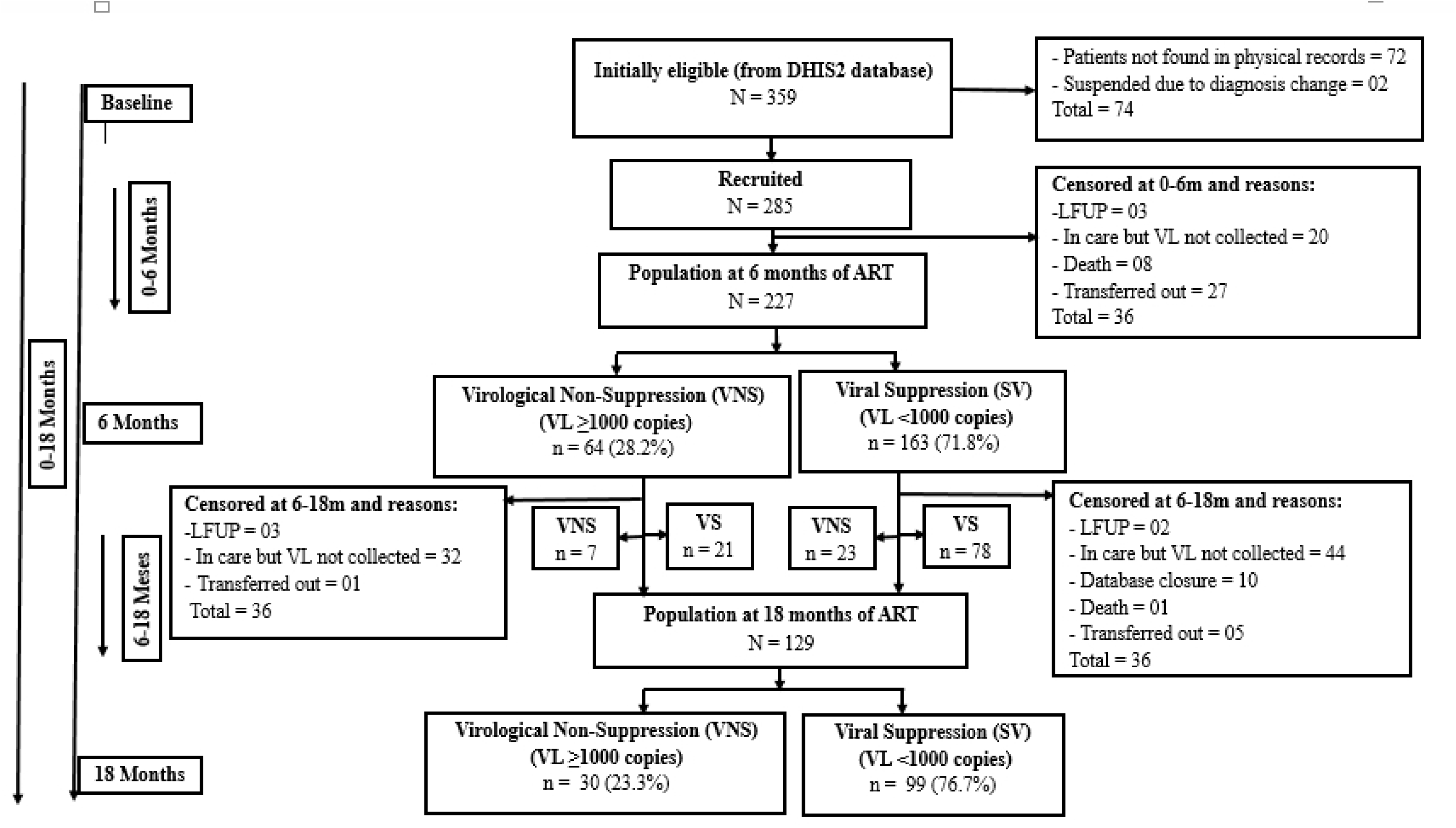
Viral load cascade and proportion of outcomes for children enrolled for the study. Chókwè Cohort Study, 2022-2024. Flowchart illustrating the VL testing cascade and virological outcomes among children <15 years receiving ART in Chókwè District, Mozambique, between 2022 and 2024. The figure presents the number of children included at 6 and 18 months of follow-up, those with available VL results, and those censored due to loss to follow-up, transfer out, death, or missing VL measurements. Among children with available VL results, the proportions achieving viral VS and experiencing VNS are shown at each time point. ART: antiretroviral therapy; VL: viral load; VS: viral suppression, VL <1000 copies/mL; VNS: virological non-suppression, VL ≥1000 copies/mL; LTFU: loss to follow-up.

### Baseline and six-month follow-up characteristics

Table 1 summarizes the characteristics of participants assessed at baseline and after 6 months of follow-up. At the baseline (ART initiation), most children were younger than five years old (median age: 3.3 years, the majority had mothers alive, presented adequate nutritional status, and were classified in WHO clinical stages I–II. Same-day ART initiation was highly frequent, and ABC/3TC + DTG was the most commonly prescribed regimen .

At 6-month follow-up, most children maintained adequate nutritional status and remained in less advanced WHO clinical stages. Good adherence to ART was observed in the majority of participants. However, psychosocial barriers affecting adherence remained common, particularly medication loss or forgetting, lack of family support, food insecurity, and transportation difficulties.

**Table 1.**
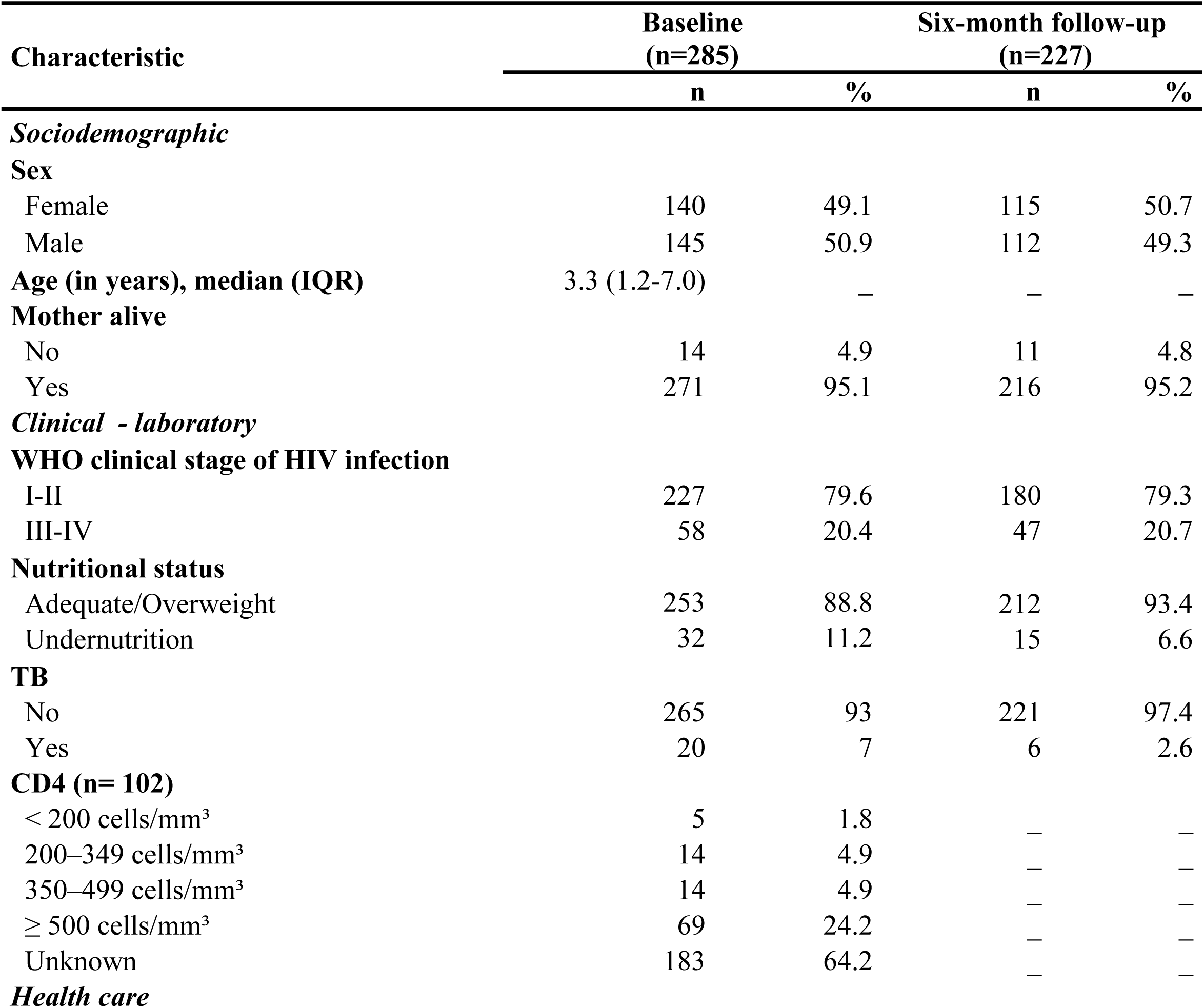
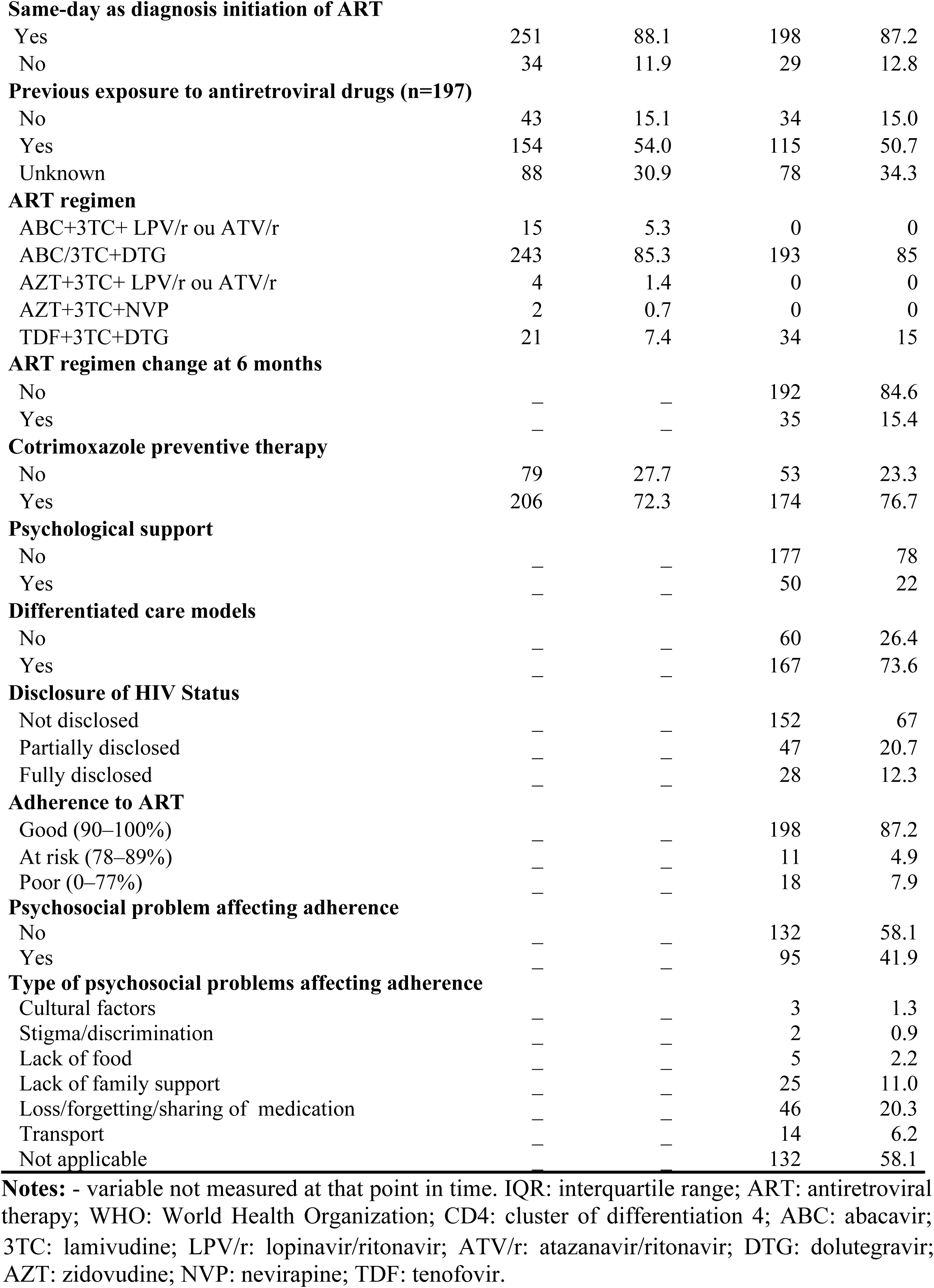
Characteristics of participants assessed at baseline and at the 6-month follow-up. Chókwè Cohort Study, 2022-2024.

| Characteristic | Baseline<br>(n=285) |  | Six-month follow-up<br>(n=227) |  |
| --- | --- | --- | --- | --- |
|  | n | % | n | % |
| ***Sociodemographic*** |  |  |  |  |
| **Sex** |  |  |  |  |
| Female | 140 | 49.1 | 115 | 50.7 |
| Male | 145 | 50.9 | 112 | 49.3 |
| **Age (in years), median (IQR)** | 3.3 (1.2-7.0) | – | – | – |
| **Mother alive** |  |  |  |  |
| No | 14 | 4.9 | 11 | 4.8 |
| Yes | 271 | 95.1 | 216 | 95.2 |
| ***Clinical - laboratory*** |  |  |  |  |
| **WHO clinical stage of HIV infection** |  |  |  |  |
| I-II | 227 | 79.6 | 180 | 79.3 |
| III-IV | 58 | 20.4 | 47 | 20.7 |
| **Nutritional status** |  |  |  |  |
| Adequate/Overweight | 253 | 88.8 | 212 | 93.4 |
| Undernutrition | 32 | 11.2 | 15 | 6.6 |
| **TB** |  |  |  |  |
| No | 265 | 93 | 221 | 97.4 |
| Yes | 20 | 7 | 6 | 2.6 |
| **CD4 (n= 102)** |  |  |  |  |
| < 200 cells/mm<sup>3</sup> | 5 | 1.8 | – | – |
| 200–349 cells/mm<sup>3</sup> | 14 | 4.9 | – | – |
| 350–499 cells/mm<sup>3</sup> | 14 | 4.9 | – | – |
| ≥ 500 cells/mm<sup>3</sup> | 69 | 24.2 | – | – |
| Unknown | 183 | 64.2 | – | – |
| ***Health care*** |  |  |  |  |
| **Same-day as diagnosis initiation of ART** |  |  |  |  |
| Yes | 251 | 88.1 | 198 | 87.2 |
| No | 34 | 11.9 | 29 | 12.8 |
| **Previous exposure to antiretroviral drugs (n=197)** |  |  |  |  |
| No | 43 | 15.1 | 34 | 15.0 |
| Yes | 154 | 54.0 | 115 | 50.7 |
| Unknown | 88 | 30.9 | 78 | 34.3 |
| **ART regimen** |  |  |  |  |
| ABC+3TC+ LPV/r ou ATV/r | 15 | 5.3 | 0 | 0 |
| ABC/3TC+DTG | 243 | 85.3 | 193 | 85 |
| AZT+3TC+ LPV/r ou ATV/r | 4 | 1.4 | 0 | 0 |
| AZT+3TC+NVP | 2 | 0.7 | 0 | 0 |
| TDF+3TC+DTG | 21 | 7.4 | 34 | 15 |
| **ART regimen change at 6 months** |  |  |  |  |
| No | — | — | 192 | 84.6 |
| Yes | — | — | 35 | 15.4 |
| **Cotrimoxazole preventive therapy** |  |  |  |  |
| No | 79 | 27.7 | 53 | 23.3 |
| Yes | 206 | 72.3 | 174 | 76.7 |
| **Psychological support** |  |  |  |  |
| No | — | — | 177 | 78 |
| Yes | — | — | 50 | 22 |
| **Differentiated care models** |  |  |  |  |
| No | — | — | 60 | 26.4 |
| Yes | — | — | 167 | 73.6 |
| **Disclosure of HIV Status** |  |  |  |  |
| Not disclosed | — | — | 152 | 67 |
| Partially disclosed | — | — | 47 | 20.7 |
| Fully disclosed | — | — | 28 | 12.3 |
| **Adherence to ART** |  |  |  |  |
| Good (90–100%) | — | — | 198 | 87.2 |
| At risk (78–89%) | — | — | 11 | 4.9 |
| Poor (0–77%) | — | — | 18 | 7.9 |
| **Psychosocial problem affecting adherence** |  |  |  |  |
| No | — | — | 132 | 58.1 |
| Yes | — | — | 95 | 41.9 |
| **Type of psychosocial problems affecting adherence** |  |  |  |  |
| Cultural factors | — | — | 3 | 1.3 |
| Stigma/discrimination | — | — | 2 | 0.9 |
| Lack of food | — | — | 5 | 2.2 |
| Lack of family support | — | — | 25 | 11.0 |
| Loss/forgetting/sharing of medication | — | — | 46 | 20.3 |
| Transport | — | — | 14 | 6.2 |
| Not applicable | — | — | 132 | 58.1 |

**Notes:** - variable not measured at that point in time. IQR: interquartile range; ART: antiretroviral therapy; WHO: World Health Organization; CD4: cluster of differentiation 4; ABC: abacavir;
3TC: lamivudine; LPV/r: lopinavir/ritonavir; ATV/r: atazanavir/ritonavir; DTG: dolutegravir; AZT: zidovudine; NVP: nevirapine; TDF: tenofovir.

### Kaplan–Meier curves of time to VNS

Kaplan–Meier survival curves were constructed to explore differences in time to VNS according to the study variables. Overall survival was first assessed for the full cohort, during which participants contributed 1,694.3 person-months with 66 events at 6 months and 3,146 person-months with 30 events at 18 months. The probability of remaining free from VNS was 31.7% (95% CI: 16.8–59.7) at 6 months and 47.3% (95% CI: 30.4–73.6) at 18 months. Significant differences in VNS-free survival were observed according to tuberculosis status at 6 months of ART, while no statistically significant differences were observed for the remaining variables (S1–S5 Figs.).

### Determinants of VNS at 6 months

In the multi-covariate 6-month ART Cox model, a 1-year increase in age at ART initiation was associated with an approximately 8% reduction in the risk of VNS Furthermore, not initiating ART on the same day as diagnosis was significantly associated with a higher risk of VNS, corresponding to an approximately 2.7-fold increase in risk (Table 2). The model was adjusted for sex, maternal vital status, baseline nutritional status, tuberculosis at ART initiation, cotrimoxazole preventive therapy, and baseline WHO clinical stage of HIV infection as a stratification variable.

**Table 2.** Cox proportional hazards models for determinants of VNS in 285 children <15 years at 6 months of follow-up. Chókwè Cohort Study, 2022-2024.

| Characteristic | Category | Crude HR |  | Adjusted HR (aHR)* |  |
| --- | --- | --- | --- | --- | --- |
|  |  | (95% CI) | p value | (95% CI) | p value |
| Sex | Male | Ref | – | Ref | – |
|  | Female | 0.96 (0.59–1.56) | 0.872 | 0.93 (0.57–1.52) | 0.766 |
| Age at ART initiation (years) | Continuous | 0.92 (0.85–1.00) | 0.053 | **0.92 (0.85–1.00)** | **0.040** |
| Mother alive | No | Ref | – | Ref | – |
|  | Yes | 0.78 (0.28–2.15) | 0.625 | 0.76 (0.27–2.13) | 0.597 |
| Baseline nutritional status | Adequate/Overweight | Ref | – | Ref | – |
|  | Undernutrition | 0.69 (0.28–1.72) | 0.425 | 0.79 (0.29–2.12) | 0.633 |
| Tuberculosis at ART initiation | No | Ref | – | Ref | – |
|  | Yes | 0.80 (0.32–2.01) | 0.640 | 0.39 (0.11–1.39) | 0.147 |
| Same day as diagnosis initiation of ART | Yes | Ref | – | Ref | – |
|  | No | 1.52 (0.81–2.84) | 0.192 | **2.67 (1.13–6.27)** | **0.024** |
| Cotrimoxazole preventive therapy at baseline | Yes | Ref | – | Ref | – |
|  | No | 0.78 (0.42–1.43) | 0.415 | 0.56 (0.29–1.10) | 0.092 |

**Notes:** Variables were selected a priori based on epidemiological relevance, as informed by previous literature and the study conceptual framework. \*Baseline clinical stage of HIV infection was included as a stratification variable. ART: antiretroviral therapy.

### Determinants of VNS at 18 months

In the multi-covariate 18-month ART Cox model, age at ART initiation showed evidence of a non-linear association with VNS. The estimated effect remained relatively constant during early childhood, with little variation in the hazard of VNS up to approximately 6 years of age. Thereafter, the hazard increased progressively with increasing age, indicating that children initiating ART at older ages were at greater risk of VNS (S6 Fig.). In addition, TB co-infection at 6 months and undernutrition at 6 months were associated with an increased risk of VNS (Table 3). The model was adjusted for sex, maternal vital status, time to ART initiation, cotrimoxazole preventive therapy, nutritional status at 6 months, TB at 6 months, psychological support, differentiated care models, adherence to ART, disclosure of HIV status, and baseline WHO clinical stage as a stratification variable.

**Table 3.**
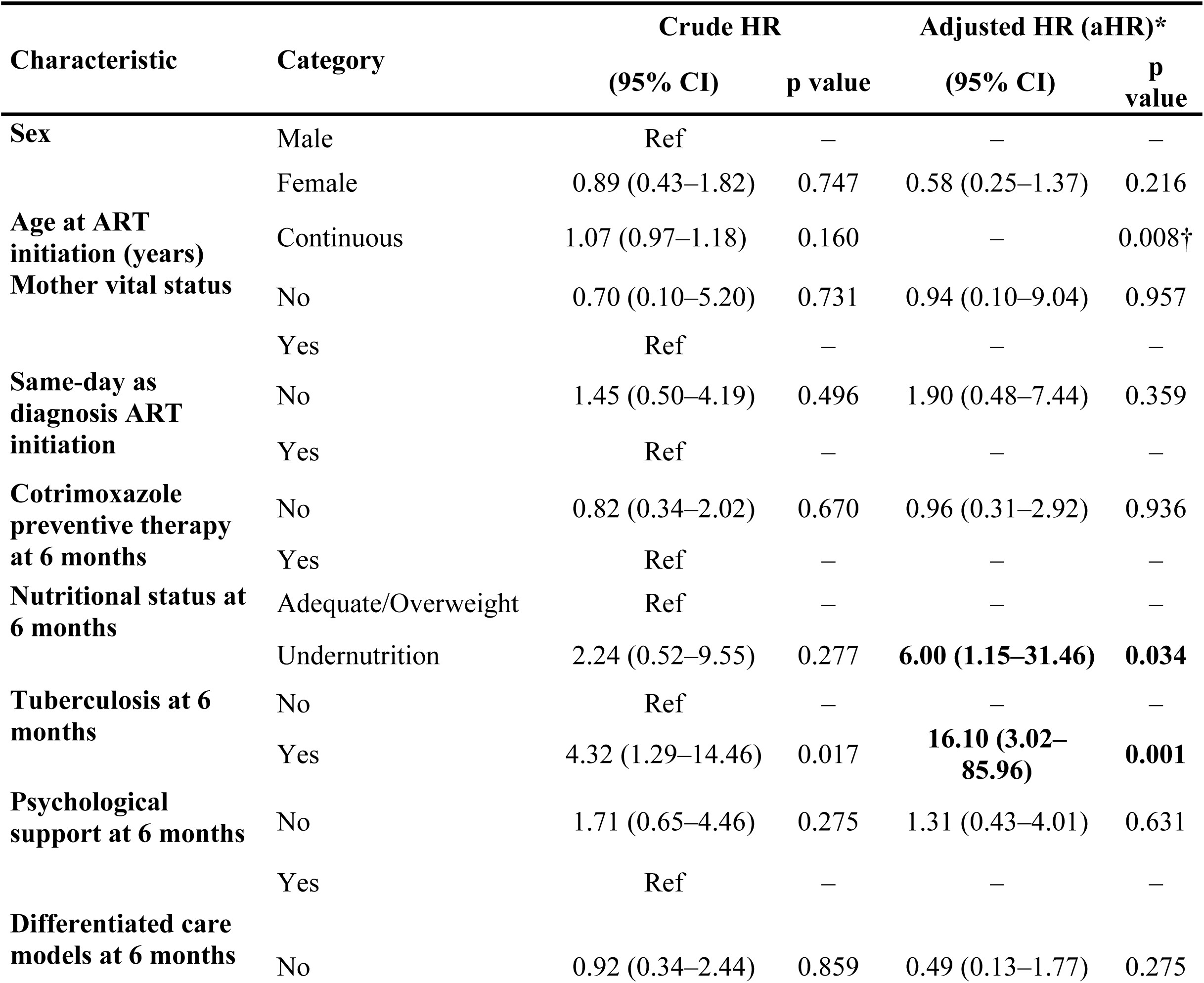
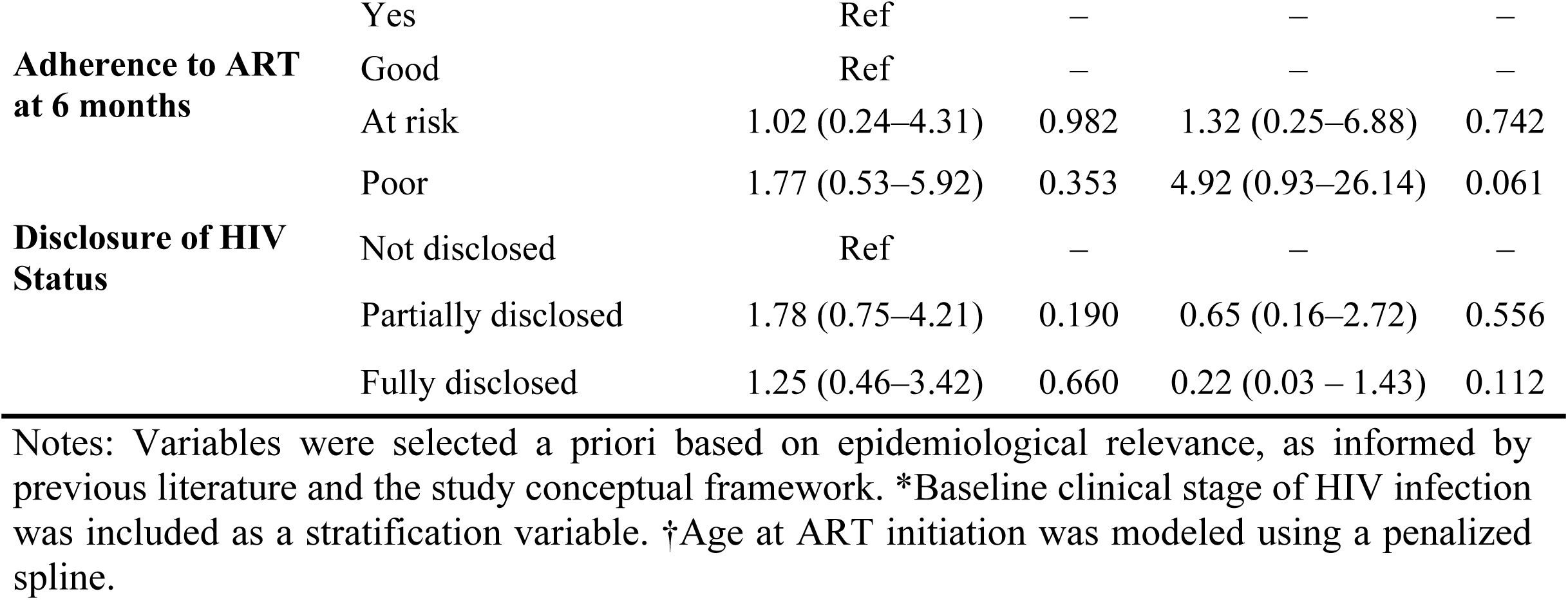
Cox proportional hazards models for determinants of VNS in 227 children under 15 years at 18 months of follow-up. Chókwè Cohort Study, 2022-2024.

## Discussion

This study analysed factors associated with VNS among children <15 years receiving ART in Chókwè District, Mozambique, after 6 and 18 months of follow-up. The results show that VNS remains frequent, occurring in 28.2% children at 6 months and in 23.3% at 18 months after ART initiation. VNS was significantly associated with younger age and late initiation of ART, as well as with TB co-infection and undernutrition.

The predominance of young children with a median age of 3.3 years, is consistent with findings from other pediatric HIV cohorts in sub-Saharan Africa, as reported in large pediatric cohorts in South Africa, where most children initiated ART before five years of age (24), and in Tanzania, where the median age was 4.3 years (25). This age distribution likely reflects increased identification and enrollment of HIV-infected children into care at younger ages following the expansion of early infant diagnosis and pediatric HIV treatment programmes.

The high proportion of children classified in WHO clinical stages I–II (79.6% at the ART initiation and 79.3% at 6 months of ART), together with the high prevalence of adequate nutritional status (88.8% at the start of ART and 93.4% at 6 months of ART), suggests relatively early diagnosis and timely initiation of ART, which may favor VS.

However, the high frequency of psychosocial problems affecting adherence, observed in approximately 42% of children at 6 months, underscores the vulnerability of this population. Previous studies have shown that barriers such as adherence difficulties, caregiver dependence, food insecurity, stigma, and structural constraints are among the main determinants of VNS in pediatric populations (26–28). These challenges remain relevant even in settings where modern pDTG-based regimens are widely available.

Evidence from Mozambique also indicates that gaps in clinical documentation, weight monitoring, and VL testing during ART implementation may indirectly worsen adherence challenges and compromise virological outcomes (29).

Our estimates of VNS are high and consistent with previous evidences from Sub-Saharan African countries. A retrospective cohort study conducted in South Africa, also reported virological failure rates of 26.5% of children experienced virological failure at 12 months treatment (24).

Similarly, a study conducted in Tanzania examining antiretroviral resistance and treatment failure among children and adolescents found that 25.4% experienced virological failure at 12 months of ART (25). In Uganda, another study investigating predictors of long-term viral failure in children and adults on ART reported that 26% of children experienced virological failure at 12 months of ART (30). A cross-sectional study carried out in Mozambique in 2019, using records of VL samples reported that only 44% (56% of VNS) of HIV-infected children <15 years achieved VS (31).

Although all results converge on the failure to meet VS targets, our results showed lower VS rates than those observed in Mozambique in 2019, as well as lower VS in the 18-month analysis compared to the six-month analysis, suggesting improvements in ART adherence in Chókwè.

Even so, VNS in the district’s pediatric population remains concerning, particularly during the first months after ART initiation. Recent evidence from Mozambique shows that, despite the national rollout of pediatric Dolutegravir (pDTG), VS among children remains suboptimal, with persistent programmatic gaps, particularly during the early phases of treatment. Approximately one-quarter of children remained with VNS after five months of follow-up, and most children eligible for pDTG initiation were younger than five years, highlighting both the challenges in achieving optimal virological outcomes and the importance of early-life HIV care within the national epidemic context (29).

Considering global VS targets, which emphasize the need to achieve high levels of VS among people on ART (1), our findings suggest that VNS continues to represent an important challenge in the local pediatric context, despite the reduction observed during follow-up. Together, these findings highlight persistent challenges in achieving and maintaining VS among children living with HIV, particularly when compared to adults, even in contexts where access to ART has expanded.

At 6 months of follow-up, age was inversely and significantly associated with VNS in the adjusted model, indicating that younger children have greater vulnerability to VNS compared to older children.

This result is consistent with findings from Mozambique (31) and others sub-Saharan African countries such as Uganda (32), Zimbabwe (33), and Tanzania (25), which also report higher risk of VNS among younger children. Several factors may explain this association, including challenges related to the palatability of pediatric formulations, frequent dose adjustments, greater clinical vulnerability, and the complete dependence on the caregiver for medication administration (27).

Furthermore, delayed initiation of ART was associated with an increased risk of VNS. This finding reinforces the importance of immediate ART initiation following diagnosis, in the line with “test-and-start” strategy, which is based on the principle that early treatment initiation improves clinical and virological outcomes (34). Evidence from African settings also indicates that delays in treatment initiation and weaknesses in the HIV care cascade are associated with poorer virological outcomes in children (26,30). These findings suggest that programmatic delays in initiating treatment may compromise therapeutic effectiveness, particularly among pediatric populations.

On the other hand, sex, maternal vital status, baseline nutritional status, TB infection at the start of ART, and the use of cotrimoxazole prophylaxis were not significantly associated with VNS at 6 months of ART. Although some studies have reported a higher risk of VNS or virological failure among male children (24,35,36), death of mother (24), malnutrition (37), TB infection (24,38,39), and those not receiving cotrimoxazole prophylaxis (40). The absence of such associations in this study may be explained by differences in sample size, statistical power, age distribution, timing of ART initiation, and ART regimens across study settings. Additionally, the widespread implementation of DTG-based regimens and same-day ART initiation in our cohort may have attenuated the effect of some traditional risk factors previously associated with virological failure.

In the 18-month adjusted model, age at ART initiation showed a significant non-linear association with VNS. Unlike the 6-month analysis, the relationship between age and VNS could not be adequately described by a simple linear effect, suggesting that the influence of age on long-term virological outcomes may vary across childhood. However, given the limited number of events and the wider confidence intervals at older ages, this finding should be interpreted with caution and confirmed in larger longitudinal studies.

At 18 months of follow-up, TB co-infection at 6 months was significantly associated with VNS in the adjusted model. However, this finding should be interpreted with caution, since only six children presented TB at this time point, resulting in a wide confidence interval and limited precision of the estimate. Similar associations have been reported in pediatric HIV studies from sub-Saharan Africa, including South Africa (24), Ghana (38), and Kenya (39). However, differences in study design, analytical approaches, and adjustment strategies may limit direct comparability across studies.

The observed association is biologically plausible. TB co-infection may increase treatment complexity, compromise adherence, and interfere with immune recovery, thereby reducing the likelihood of sustained virological suppression (41). In addition, co-infection may contribute to the development of drug resistance, particularly when clinical and virological monitoring is insufficient (42). In Mozambique, where TB co-infection remains a major public health challenge, strengthening the integration of TB and HIV services may help improve long-term virological outcomes among children (29).

Furthermore, undernutrition at 6 months of ART was associated with an increased risk of VNS at 18 months of follow-up. Children with undernutrition had a sixfold higher risk of VNS compared with those with overweight/adequate nutritional status after adjustment with other relevant covariates.

The observed association is biologically plausible. Undernutrition may impair immune recovery, increase susceptibility to opportunistic infections, and compromise the absorption and metabolism of antiretroviral drugs, potentially reducing treatment effectiveness. Previous studies among children living with HIV in sub-Saharan Africa have shown that undernutrition is associated with poorer clinical outcomes, delayed immune recovery, and increased mortality among children receiving ART, highlighting the persistent interaction between nutritional status and HIV disease progression (37,43,44). In addition, undernutrition often reflects broader socioeconomic vulnerabilities, including food insecurity, poverty, and limited caregiver resources, which may negatively affect adherence and continuity of care. In resource-limited settings such as Mozambique, where HIV and undernutrition commonly coexist, these biological and social mechanisms may act synergistically to compromise long-term virological outcomes among children receiving ART.

Other variables did not independently influence VNS at 18 months of ART in this cohort. However, other studies reported that poor adherence to ART (11), and non-disclosure of HIV status to the child (40,45), were independently associated with this outcome. The lack of association observed in this study may be explained by the predominance of very young children in the cohort, whose treatment adherence depends largely on caregivers. Previous studies also suggest that HIV status disclosure has a greater impact on treatment adherence among older children and adolescents (46).

This study has some limitations that should be considered when interpreting the findings. First, the retrospective design, based on routinely collected secondary data from health services, may have affected data accuracy and completeness, potentially introducing information and measurement bias. However, the use of routinely collected programmatic data enhances the external validity of the findings, as the study reflects real-world pediatric HIV care conditions in a high-burden district of Mozambique.

Second, a substantial proportion of missing data (reported as “unknown” in the dataset), particularly for CD4 count (64.2%) and previous antiretroviral exposure (30.9%), limited their inclusion in the multivariable analyses. The high proportion of missing CD4 values likely reflects operational challenges in routine pediatric HIV care and laboratory monitoring in resource-limited settings. Nevertheless, key variables related to the study objectives, including VL outcomes, nutritional status, tuberculosis status, and ART-related characteristics, were available for most participants and allowed robust assessment of the main determinants of VNS.

Additionally, attrition during follow-up should be considered when interpreting the 18-month findings, since VL results were available for a smaller subset of participants at this time point. This may have introduced selection bias or survivor bias, as children retained in care and with available VL measurements may differ systematically from those lost to follow-up or without laboratory monitoring. However, the longitudinal cohort design and the use of survival analysis methods, particularly Cox regression, allowed the appropriate handling of censored observations and maximized the use of available follow-up time, strengthening the validity of the estimates despite incomplete follow-up.

Finally, despite efforts to ensure data quality through cross-checking of multiple data sources (patients’ paper charts and electronic medical records), residual misclassification cannot be ruled out. Nonetheless, the use of multiple data sources likely reduced recording errors and improved data completeness compared with reliance on a single source of information.

Despite these limitations, this study provides important evidence on determinants of VNS among children receiving ART in Mozambique. To our knowledge, it is one of the few longitudinal studies in the country to evaluate factors associated with VNS at both 6 and 18 months of follow-up using time-to-event methods, generating findings with direct relevance for pediatric HIV programs in similar resource-limited settings.

### Programmatic and public health implications

Overall, the findings of our study support regional evidence indicating that VNS among children is multifactorial, resulting from the interaction of age at ART initiation, TB co-infection and undernutrition. These results highlight the need for integrated pediatric HIV care strategies that address both early-life vulnerability and clinical and nutritional comorbidities throughout the continuum of care.

From a programmatic perspective, our results suggest the need to strengthen routine pediatric HIV care through targeted and practical interventions. These include prioritizing same-day ART initiation, as well as implementing enhanced follow-up strategies during the first months of treatment, particularly for younger children. More frequent clinical and adherence monitoring during this early period–such as monthly follow-up visits or differentiated care models with closer supervision–may help identify adherence barriers earlier and prevent treatment failure.

In addition, active patient tracing mechanisms, including home visits and community-based follow-up by community health workers, could play a key role in reducing loss to follow-up and improving retention in care. Strengthening caregiver-centered interventions, such as structured adherence counseling, psychosocial support, and education on pediatric ART management, is also essential, given children’s dependence on caregivers for treatment adherence.

Furthermore, our findings highlight the importance of integrating TB, HIV, and nutritional services within routine pediatric care. Systematic TB screening, prompt management of co-infection, regular nutritional assessment, and early identification of growth faltering should be prioritized, particularly among younger children. Notably, undernutrition emerged as an independent predictor of VNS at 18 months of follow-up, highlighting the importance of nutritional status not only as an indicator of child health but also as a determinant of long-term virological outcomes among children receiving ART. Expanding access to nutritional support interventions, differentiated service delivery models tailored for children, and routine VL monitoring with timely clinical decision-making may further enhance treatment success.

Together, these strategies may improve adherence, reduce early treatment failure, and accelerate progress toward achieving the 95–95–95 targets in pediatric HIV care in resource-limited settings.

## Conclusion

This study highlights that VNS remains an important challenge among children receiving ART in Chókwè District, despite improvements in virological outcomes. The findings suggest that sustained VS is influenced by the interplay of clinical, nutritional, age-related, and programmatic factors throughout the continuum of care. Strengthening early HIV diagnosis, timely ART initiation, integrated management of TB and malnutrition, and providing individualized age-responsive follow-up throughout childhood may improve treatment outcomes and accelerate progress toward achieving the UNAIDS 95–95–95 targets among children living with HIV in resource-limited settings.

## Data Availability

The de-identified dataset underlying the findings of this study is publicly available in Zenodo at https://doi.org/10.5281/zenodo.21242355

## Acknowledgements

The authors would like to thank the Chókwè District Health Service, the Gaza Provincial Health Directorate, and all health professionals involved in pediatric HIV care in Chókwè for their support and collaboration. We also acknowledge the teams responsible for data management and health information recording in the facilities, whose work made this study possible.

We are especially grateful to the children and their caregivers whose data contributed to this study. Although collected through routine clinical care and anonymized records, their information represents an invaluable contribution to scientific knowledge and to the improvement of HIV services in Mozambique.

## Supporting information

**S1 Table. Domain, operational definitions, and categorization of variables used in the Cox regression analyses of VNS among children <15 years receiving ART. Chókwè Cohort Study, 2022-2024.** Variables were obtained from routine clinical recordsand electronic medical records. VNS was defined according to the Mozambican national HIV guidelines as a viral load ≥1,000 copies/mL after at least six months of ART.

**S1 Fig. Kaplan–Meier curves for time to VNS at 6 months of follow-up according to baseline characteristics among children <15 years receiving. Chókwè Cohort Study, 2022-2024.** Panel A: age ; Panel B: sex; Panel C: mother vital status; and Panel D: same-day ART initiation.

**S2 Fig. Kaplan–Meier curves for time to VNS at 6 months of follow-up according to baseline characteristics among children <15 years receiving ART. Chókwè Cohort Study, 2022-2024.** Panel E: cotrimoxazole preventive therapy; Panel F: tuberculosis at ART initiation; Panel G: nutritional status; and Panel H: WHO clinical stage.

**S3 Fig. Kaplan–Meier curves for time to VNS at 18 months of follow-up according to 6 months characteristics among children <15 years receiving ART. Chókwè Cohort Study, 2022-2024.** Panel I: sex; Panel J: mother vital status; Panel K: starting ART at the same-day of diagnostic; and Panel L: cotrimoxazole preventive therapy.

**S4 Fig. Kaplan–Meier curves for time to VNS at 18 months of follow-up according to 6 months characteristics among children <15 years receiving ART. Chókwè Cohort Study, 2022-2024.** Panel M: TB co-infection; Panel N: nutritional status; Panel O: WHO clinical stage of HIV infection; and Panel P: psychological support.

**S5 Fig. Kaplan–Meier curves for time to VNS at 18 months of follow-up according to 6 months characteristics among children <15 years receiving ART. Chókwè Cohort Study, 2022-2024.** Panel Q: differentiated care models; Panel R: adherence to ART; and Panel S: disclosure of HIV status.

**S6 Fig. Adjusted non-linear association between age at ART initiation and the risk of virological non-suppression at 18 months of follow-up. Chókwè Cohort Study, 2022–2024.** Estimated partial effect of age at ART initiation on the hazard of virological non-suppression derived from a multi-covariate Cox proportional hazards model using penalized splines. The model was adjusted for sex, maternal vital status, same-day ART initiation, cotrimoxazole preventive therapy at 6 months, nutritional status at 6 months, tuberculosis at 6 months, psychological support, differentiated care models, adherence to ART, and disclosure of HIV status, with baseline WHO clinical stage included as a stratification variable. The solid line represents the estimated log hazard ratio, and the dashed lines represent the 95% confidence intervals. The estimated hazard remained relatively stable during early childhood and increased progressively from approximately 6 years of age onward.

